# An Evaluation of the Safety and Immunogenicity of MVC-COV1901: Results of an interim analysis of a phase III, parallel group, randomized, double-blind, active-controlled study

**DOI:** 10.1101/2022.07.14.22277617

**Authors:** Julio Torales, Osmar Cuenca-Torres, Laurentino Barrios, Luis Armoa- Garcia, Gladys Estigarribia, Gabriela Sanabria, Meei-Yun Lin, Josue Antonio Estrada, Lila Estephan, Hao-Yuan Cheng, Charles Chen, Robert Janssen, Chia-En Lien

## Abstract

**Background:** Data from previous studies of the MVC-COV1901 vaccine, a subunit vaccine against SARS-CoV-2 based on the stable prefusion spike protein (S-2P) adjuvanted with CpG 1018 adjuvant and aluminum hydroxide, suggest that the vaccine is generally safe and elicits a good immune response in healthy adults and adolescents. By comparing with AZD1222, this study adds to the findings from previous trials and further evaluates the breadth of protection offered by MVC-COV1901.

**Methods:** In this phase 3, parallel group, randomized, double-blind, active-controlled trial conducted in 2 sites in Paraguay, we assigned adults aged 18 to 91 years in a 1:1 ratio to receive intramuscular doses of MVC-COV1901 or AZD1222 administered as scheduled in the clinical trial. Serum samples were collected on the day of vaccination and 14 days after the second dose. Primary and secondary safety and immunogenicity endpoints were assessed. In addition, other outcomes investigated were cross-reactive immunity against the Omicron strain and the induction of IgG subclasses.

**Results:** A total of 1,030 participants underwent randomization. Safety data was derived from this set while primary immunogenicity data involved a per-protocol immunogenicity (PPI) subset including 225 participants. Among the participants, 58% are seropositive at baseline. When compared against AZD1222, MVC-COV1901 exhibited superiority in terms of neutralizing antibody titers and non-inferiority in terms of seroconversion rates. Reactogenicity was generally mild and no serious adverse event was attributable to MVC-COV1901. Both vaccines have a Th1-biased response predominated by the production of IgG1 and IgG3 subclasses. Omicron-neutralizing titers were 44.5 times lower compared to wildtype-neutralizing titers among seronegative individuals at baseline. This fold-reduction was 3.0 times among the seropositive.

**Conclusion:** Results presented here demonstrate the safe and robust immunogenicity from MVC-COV1901. Previous infection coupled with vaccination of this vaccine may offer protection against the Omicron strain though its durability is still unknown.

**ClinicalTrials.gov registration:** NCT05011526

## Introduction

Since it was first reported in December 2019, COVID-19 has rapidly spread affecting millions of lives and causing over 6 million deaths worldwide [1]. High transmission rates have threatened and quickly overwhelmed health systems. The urgency of addressing this pandemic has led to the accelerated development of vaccines from various platforms. To date, over 63.4% of the world’s population has received at least one dose of a COVID-19 vaccine but the inequitable distribution of vaccine remains a problem. It is estimated that only 13.6% of people in low-income countries have received at least one dose of an approved COVID-19 vaccine [1]. Further increasing vaccine coverage and improving vaccine equity require surpassing logistical and supply constraints and the use of vaccines that are proven safe and effective and offer breadth in terms of protection

During late 2020, reports from different countries confirmed the emergence of SARS-CoV-2 variants that caused differing degrees of transmission, ability to cause severe disease and immune evasion [2]. In November 2021, the first case of the Omicron variant was reported in South Africa [3]. Although found to have a lesser risk for severe disease than the previously discovered Delta variant, mutations in the Omicron equipped the variant with greater transmissibility, effectivity in avoiding the human immune response and resistance against some existing treatments [4,5]. By the end of March 2022, more than 90% of SARS-CoV-2 infections worldwide were caused by this variant [1]. For populations receiving insufficient protection, an epidemic of the virus may lead to detrimental effects. Although the incidence of severe disease is lower, higher transmission leading to a large volume of cases, could overwhelm health systems and threaten economies. As aforementioned, it is crucial that we find solutions that are safe and effective and offer protection against the different variants of the virus.

MVC-COV1901 is a subunit vaccine based on the stable prefusion spike protein (S-2P) of SARS-Cov-2 adjuvanted with CpG 1018 adjuvant and aluminum hydroxide and has been approved for use in 3 countries [6,7]. A large phase 2 clinical trial has demonstrated its favorable safety and immunogenicity profiles [8]. An immunobridging assessment demonstrated non-inferiority in terms of immunogenicity to AZD1222 with a geometric mean titer (GMT) of neutralizing antibodies equivalent to 3.8 times that of AZD1222. [9]. In terms of safety, the V-Watch program, launched by the Taiwan Centers for Disease Control to monitor post-marketing safety, reported no serious adverse effects for MVC in 2 million doses administered [10]. Among adolescents, the vaccine was well tolerated and had an immunogenic effect that is non-inferior to that of young adults [11]. For stability, this vaccine can be transported and stored in standard refrigeration temperatures and can, therefore, be easily utilized in low-resource settings [12].

Here, we report an interim analysis of a phase 3 trial to assess the safety, tolerability, and immunogenicity of two doses of the MVC-COV1901 compared with AZD1222. We assessed the superiority of MVC-COV1901 against AZD1222 in terms of neutralizing antibodies and its non-inferiority in terms of seroconversion. We also looked at other dimensions of immunity particularly the profile of IgG subclass antibody responses among vaccinated individuals. As a huge majority of the infections globally are caused by the Omicron variant, it is imperative that we investigate the immunogenicity of the vaccine against Omicron. This ongoing trial, which began in October 2021, was conducted at the time the Omicron variant was widely circulating. We, therefore evaluated the immune response induced by two doses of the vaccine against the Omicron (BA.1) strain in both the seropositive and seronegative subsamples at baseline.

Seropositive individuals considered in this analysis were those with possible prior infection of COVID-19 as indicated by a reactive anti-N protein test and pre-vaccination anti-spike antibody titers that are above the lower limit of detection.

## Methods

### Study design and participants

The MVC-COV1901 phase 3 trial was a parallel-group, prospective, randomized, double-blind, active-controlled, and multi-center study to evaluate the safety, tolerability, and immunogenicity of the SARS-CoV-2 vaccine candidate MVC-COV1901 compared to AZD1222 in adult volunteers of 18 years and above (NCT05011526). Figure 1 outlines the trial profile and study schematic. The main study of the trial consisted of 1,030 subjects ≥ 18 years of age who were generally healthy or with stable pre-existing medical conditions recruited from two study sites - Asuncion and Ciudad del Este-in Paraguay. A full list of inclusion and exclusion criteria can be found in the appendix. This sample of 1,030 individuals was used for safety analysis. From this sample, the immunogenicity subset which consisted of 884 subjects, was derived. The per-protocol immunogenicity (PPI) analysis set included 225 participants.

**Figure 1.**
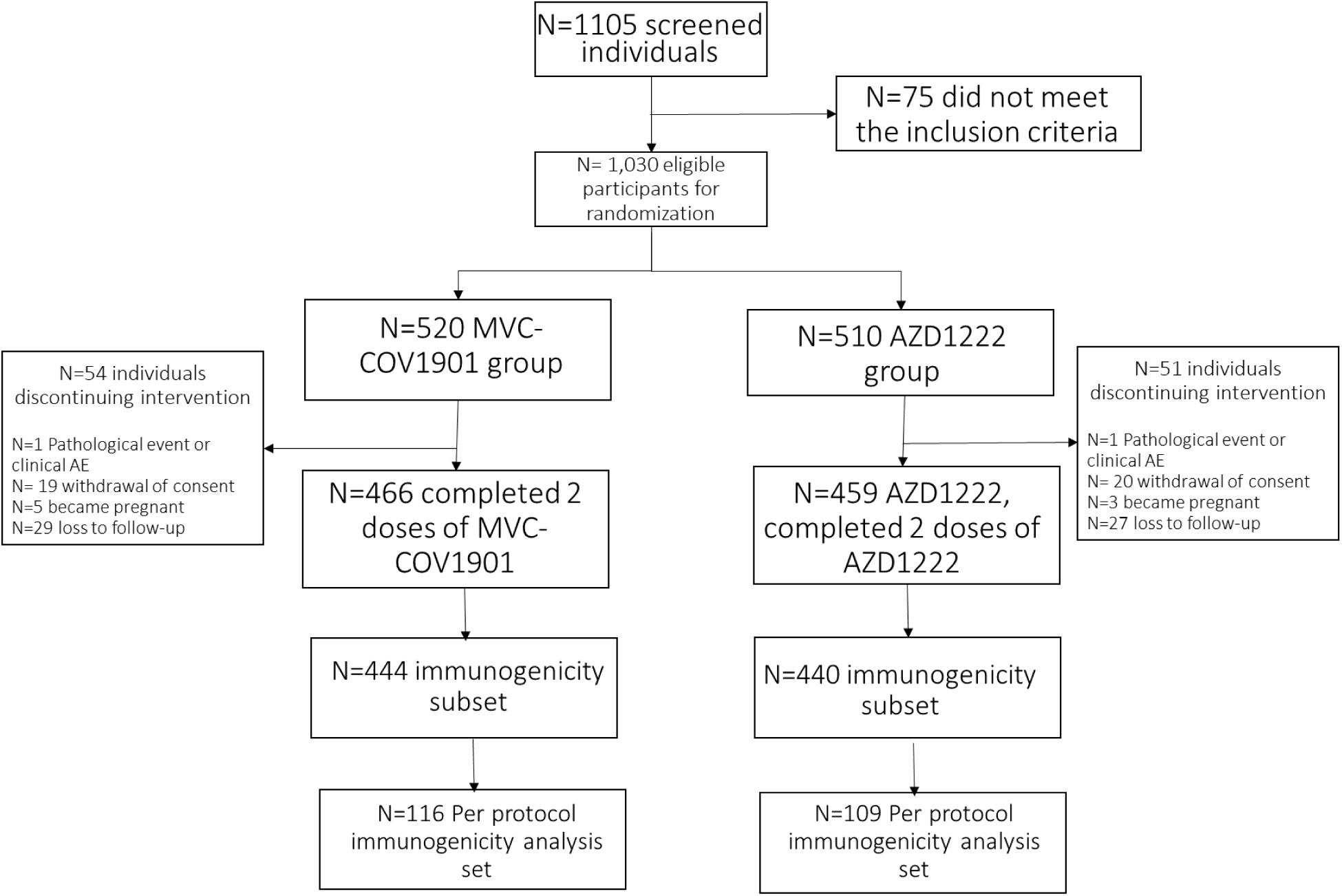
Trial profile

The trial protocol and informed consent form were approved by the local regulatory entity (DINAVISA) and the ethics committees at the participating sites. The main institutional review board was the National University of Asuncion. The phase 3 trial was done in accordance with the principles of the Declaration of Helsinki.

### Randomization and blinding

Eligible participants were unvaccinated individuals randomized to receive either MVC-COV1901 or AZD1222 in a 1:1 ratio. All participants were randomly assigned to study intervention using an interactive web response system (IWRS). Simple random sampling was used to select study participants by study site.

Double-blinding was employed in the study; hence, both participants and investigators were blinded to the participants’ assignment of the study intervention. In case of emergency, the investigator held the sole responsibility of determining if unblinding of intervention assignment is warranted. Once Day 43 (or 14 days after the second dose) was reached, the interim analysis was carried out. Participants, investigators, site personnel, local regulators, and the sponsor staff were then unblinded to the treatment group assignments.

### Procedures

Subjects received two doses of either MVC-COV1901 or AZD1222 as scheduled in the clinical trial. Both vaccines were delivered intramuscularly at the deltoid. Serum samples were collected on the day of vaccination (Day 1) and 14 days after the second dose (Day 43 after the first dose).

The Medigen COVID-19 vaccine, MVC-COV1901, is a subunit vaccine consisting of the prefusion spike protein (S-2P), 750 μg CpG 1018 adjuvant and 375 μg aluminum hydroxide. A standard 0.5 mL dose contains 15 µg of the Spike-2P. The active control is AZD1222, an adenoviral vector vaccine developed by Oxford University and AstraZeneca served in multi-dose vials. Each dose of vaccine is 0.5 mL and contains 5 × 10^10^ viral particles.

For safety analysis, vital signs were checked before and after each injection. Individuals injected with the intervention were observed for at least 30 minutes after administration of the intervention. This period is for the assessment of immediate adverse events. Participants who received at least one dose of the study intervention were evaluated for safety. Data from these were reported as part of the safety set. Participants were then asked to record any local or systemic adverse events for up to 7 days after each injection. Unsolicited adverse events were reported for 28 days after each dose of the vaccines. Other adverse events, serious adverse events, adverse events of special interest, and vaccine-associated enhanced diseases were noted throughout the study period. Levels of the severity of solicited and unsolicited adverse events were reported using modified grading scales from the US FDA Guidance for Industry [13]. The levels of severity were not noted for the reporting of “diarrhea” and “nausea” under solicited systemic AEs.

For immunogenicity analysis, assessments were conducted on Day 1 and Day 43. Measurement of neutralizing antibody titers was performed by central laboratories in Taiwan using validated live virus neutralization assay while determination of antigen-specific immunoglobulin (IgG) titers was performed by a laboratory in Taiwan using validated enzyme-linked immunosorbent assay (ELISA) [8]. In brief terms, the live virus neutralization assay was conducted by mixing serially-two-fold diluted sera with an equal volume of SARS-CoV-2 virus (hCoV-19/Taiwan/4/2020, GISAID accession ID: EPI_ISL_411927). This serum-virus mixture was incubated and added to Vero E6 cells before further incubation. Neutralizing antibody titer (NT50) was defined as the reciprocal of the highest dilution capable of inhibiting 50% of the cytopathic effect. The NT_50_ results were calculated with the Reed-Muench method. For antigen-specific immunoglobulin titers, serum samples were analyzed in Taiwan using the ELISA method with plates coated with S-2P proteins. GMTs obtained through the assays were converted to standardized units: IU/mL for neutralizing antibody titers and BAU/mL for antigen-specific immunoglobulin. For neutralizing antibodies, titers were converted using the equation: y = 1.5001(x^0.8745^); where *x* is the value of the GMT. For the IgG titers, titers tested in Taiwan were converted to BAU/mL by multiplying by a conversion factor of 0.0912[8]. For local analysis and usage, IgG titers were also tested in a laboratory in Paraguay. Antigen-specific immunoglobulin titers were tested using the LIAISON^®^ SARS-CoV-2 TrimericS IgG assay, a chemiluminescence immunoassay (CLIA) used for the quantitative determination of anti-trimeric spike protein-specific IgG antibodies to SARS-CoV-2 in serum or plasma samples [14].

For the evaluation of the Omicron-neutralizing ability, neutralizing assays using pseudovirus with spike proteins of Wuhan wild-type or the Omicron variant were performed as in the previous study [12]. Twofold serial dilutions of serum samples were mixed with equal volumes of pseudovirus and incubated before adding to the HEK293-hAce2 cells. Fifty percent dilution titers (ID50) were calculated considering the uninfected cells as 100% neutralization and cells transduced with the virus as 0% neutralization.

### Outcomes

The outcomes evaluated in this study were safety, tolerability, and immunogenicity. The primary safety outcomes involved the evaluation of the safety and tolerability of MVC-COV1901 versus AZD1222. Primary safety endpoints include immediate adverse events, solicited local and systemic adverse events (evaluated up to 7 days after each dose of the study intervention), and unsolicited adverse events (assessed up to 28 days after each dose of the study intervention). The primary immunogenicity outcomes, on the other hand, were measured in wild-type anti-SARS-CoV-2 virus-neutralizing antibody GMTs, GMT ratio between MVC-COV1901 and AZD1222, geometric mean fold rise (GMFR) from baseline antibody levels, and seroconversion rates (SCR) at day 14 after the second dose of the vaccine. Seroconversion is defined as at least a 4-fold increase of post-intervention antibody titers from the baseline titers or half of the lower limit of detection (LoD), if undetectable, at baseline. The study aims to determine superiority in neutralizing antibodies and non-inferiority in terms of SCR of MVC-COV1901 against AZD1222 measured 14 days after the second dose of the study intervention. Superiority is established when the lower limit of the 95% confidence interval (CI) of the GMT ratio (MVC-COV1901/AZD1222) is greater than 1, while non-inferiority in SCR is considered when the lower limit of the 95% CI of the difference in SCRs (MVC-COV1901/AZD1222) is greater than or equal to -5%.

Secondary safety outcomes assessed included serious adverse events, adverse events of special interest, and vaccine-associated enhanced diseases, while the secondary immunogenicity outcome considered was the level of antigen-specific immunoglobulin or IgG (in BAU/mL) measured at Day 43 (i.e. 14 days after the second dose).

This study also investigated other dimensions of immunity induced by the primary regimen of the MVC-COV1901. The other outcomes being investigated were cross-reactive immunity against the Omicron strain of SARS-CoV-2 and non-neutralizing antibody immune effector mechanisms particularly the induction of IgG subclasses.

### Statistical analysis

The primary safety outcomes in this interim analysis were assessed in the safety set, which, as previously mentioned, included randomly assigned participants who had at least one dose of study intervention. Primary immunogenicity endpoints were evaluated using the PPI subset consisting of participants who had received two doses of the study intervention, had valid immunogenicity data on Day 43, and had no major protocol deviations up to Day 43.

We calculated the sample size based on the following assumptions: (1) Level of significance = 0.025 (one-sided), (2) Level of power = 0.9, (3) Expected geometric mean ratio of MVC-COV1901 to AZD1222 = 1.2 and (4) SD of natural log data = 0.81 [8]. Under the above assumptions, a sample of 417 participants per group provides a power of 90% to establish superiority of MVC-COV1901 to AZD1222 in terms of GMT ratio of neutralizing antibody titers at day 43. Furthermore, we considered a dropout rate of 11.4% resulting to a total of at least 942 participants considered for the study.

All measured variables and derived parameters were listed by individual participant and analyzed using descriptive statistics. Summary descriptive statistics were provided for demographic/baseline characteristics, secondary immunogenicity, safety, exploratory immunogenicity, and efficacy variables. Continuous variables were summarized descriptively with the number of participants, mean, median, standard deviation (SD), interquartile range (IQR), range (minimum and maximum), and 95% CI of mean and median (when appropriate). Categorical variables were summarized with the number and percentage of participants. The geometric means of IgG and neutralizing antibody titers were calculated together with their 95% CIs, whereas GMT ratios, and their 95% CIs were obtained from the ratio of the GMTs of MVC and AZ. Significance tests (2-tailed, alpha = 0.05) without alpha adjustment were performed for pairwise comparison where appropriate and p-value was rounded to four decimal places as applicable.

## Results

### Demographics and baseline characteristics

Table 1 shows the demographic profile and baseline characteristics of the participants. We found that both groups are comparable across all baseline characteristics. There are no statistically significant differences between the two vaccine groups when considering the different demographic variables (p>0.05). In terms of age, both groups are similar with a mean age of 32.1 years for the MVC-COV1901 group and 32.2 years for the AZD1222 group (p=0.8929). Both groups are predominantly Latino or Hispanic and are primarily males. As in age, BMI of participants were also similar for the two groups (p=0.4318). Lastly, there are more individuals without any comorbidity in both groups.

**Table 1.** Demographic profile and baseline characteristics of participants.

| Statistic | AZD1222<br>N=510 | MVC-COV1901<br>N=520 | Total<br>N=1030 | p-value |
| --- | --- | --- | --- | --- |
| Age (years) |  |  |  |  |
| Mean (SD) | 32.2 (11.70) | 32.1 (12.12) | 32.1 (11.91) | 0.8929 |
| Median | 29.0 (16.0) | 28.0 (16.0) | 29.0 (16.0) |  |
| Min, Max (Range) | 18.0 ~ 71.0 (53) | 18.0 ~ 91.0 (73) | 18.0 ~ 91.0 (73) |  |
| Gender (n/%) |  |  |  |  |
| Male | 315 (61.8) | 304 (58.5) | 619 (60.1) | 0.279 |
| Female | 195 (38.2) | 216 (41.5) | 411 (39.9) |  |
| Ethnicity (n/%) |  |  |  |  |
| Latino or Hispanic | 501 (98.2) | 509 (97.9) | 1010 (98.1) | 1 |
| White or Caucasian | 8 (1.6) | 9 (1.7) | 17 (1.7) |  |
| Asian | 1 (0.2) | 2 (0.4) | 3 (0.3) |  |
| BMI (kg/m2) |  |  |  |  |
| Mean (SD) | 27.6 (5.90) | 27.3 (6.33) | 27.5 (6.11) | 0.432 |
| Median | 26.4 (7.7) | 26.2 (7.4) | 26.3 (7.6) |  |
| Min, Max (Range) | 17.0 ~ 54.7 | 14.9 ~ 60.1 | 14.9 ~ 60.1 |  |
| Comorbidity |  |  |  |  |
| without | 336 (65.9) | 330 (63.5) | 666(64.7) | 0.416 |
| Any | 174 (34.1) | 190 (36.5) | 364 (35.3) |  |
| Arrhythmia | 1 (0.2) | 1 (0.2) | 2 (0.2) |  |
| Arterial Hypertension | 35 (6.9) | 25 (4.8) | 60 (5.8) |  |
| Asthma | 8 (1.6) | 17 (3.3) | 25 (3.4) |  |
| Cancer | 2 (0.4) | 1 (0.2) | 3 (0.3) |  |
| Chronic Obstruction<br>Pulmonary Disease (COPD) | 0 | 4 (0.8) | 4 (0.4) |  |
| Diabetes | 12 (2.4) | 11 (2.1) | 23 (2.2) |  |
| History of drug allergy | 20 (3.9) | 13 (2.5) | 33 (3.2) |  |
| Kidney Disease | 0 | 4 (0.8) | 4 (0.4) |  |
| Liver disease | 1 (0.2) | 3 (0.6) | 4 (0.4) |  |
| Obesity | 106 (20.8) | 108 (20.8) | 214 (20.8) |  |

### Safety outcomes

The occurrence of adverse events is summarized in Figure 2 (Table S1-1 and S1-2). Overall, a total of 465 (45.1%) participants reported solicited local AEs after any dose of the study intervention: 342 (33.2%) participants after the first dose of study intervention and 239 (25.8%) participants after the second dose of study intervention. The proportion of participants who reported solicited local adverse events after first and second doses of the study intervention was slightly higher in the AZD1222 group than in the MVC-COV1901 group. Majority of these participants reported Grade 1 (mild) and some reported Grade 2 (moderate) solicited local adverse events after any dose of study intervention. Pain or tenderness and injection site pruritus was the most frequently reported solicited local AE. Grade 3 adverse events were reported for pain or tenderness, injection site pruritus, and hematoma.

**Figure 2.**
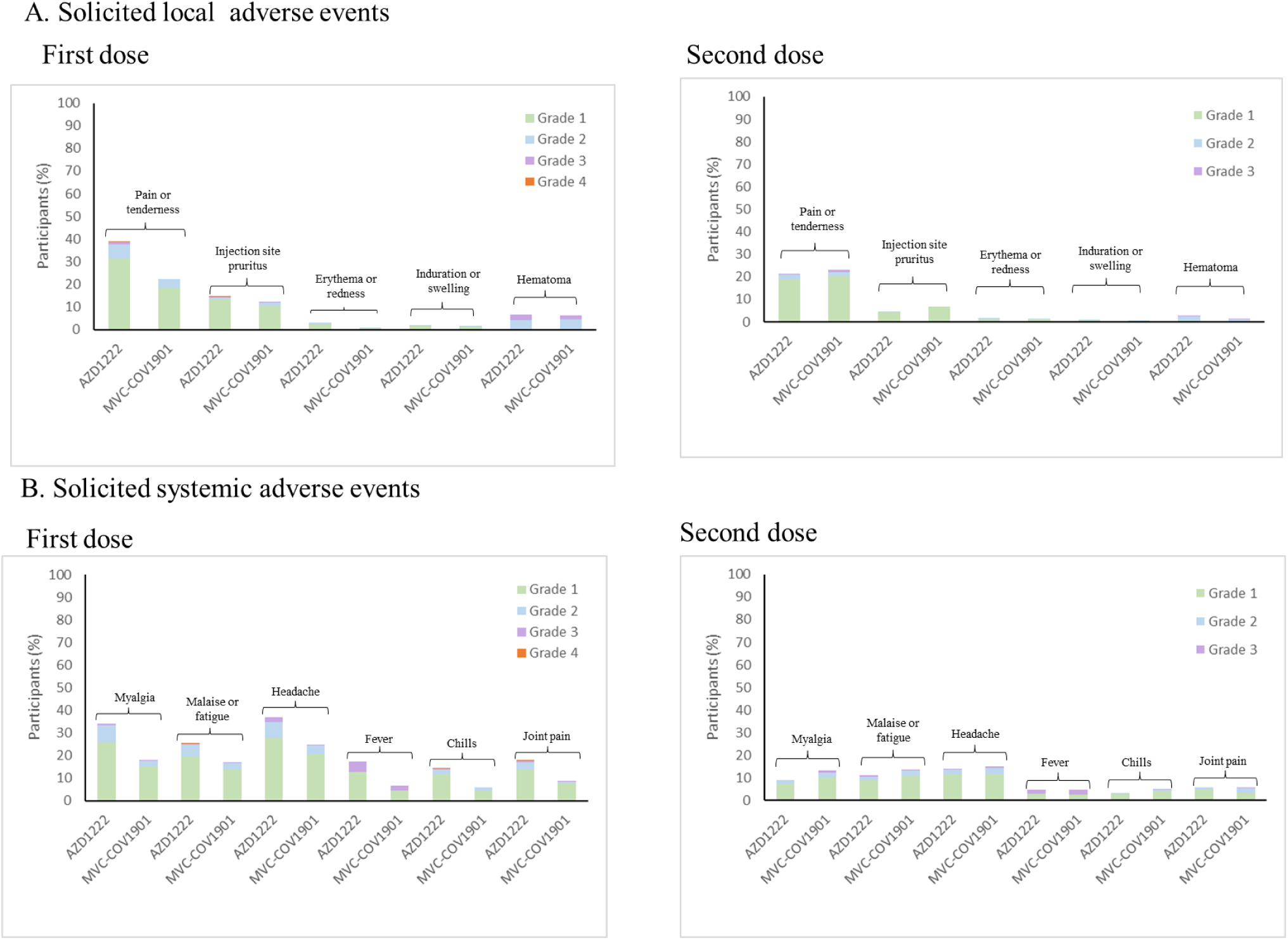
Solicited local (A) and systemic (B) adverse events occurring within 7 days of the first and second doses of MVC-COV1901 or AZD1222. Adverse events were graded as mild (grade1), moderate (grade 2), severe (grade 3), or disabling (grade 4)

A total of 552 (53.6%) participants reported solicited systemic adverse events after any dose of the study intervention: 480 (46.6%) participants after the first dose of study intervention and 222 (24.0%) participants after the second dose of study intervention. The proportion of participants who reported solicited systemic adverse events after any dose of the study intervention was slightly higher in the AZD1222 group than in the MVC-COV1901 group, specifically after the first dose of the study intervention. Most of these participants reported Grade 1 (mild) and a minority reported Grade 2 (moderate) solicited systemic adverse events. The most common solicited systemic adverse events are headache and myalgia. Grade 3 (severe) headaches were reported in the MVC-COV1901 group while a single case of Grade 4 myalgia was reported in the same group. For malaise or fatigue, 2 participants reported Grade 3 incidents after the second dose of MVC. Other reports of Grade 3 solicited systemic adverse events were made for fever, chills and joint pain.

A total of 16 (1.6%) participants reported unsolicited adverse events. The most frequently reported of these were gastrointestinal disorders (0.2%) and hypertension (0.2%). No unsolicited adverse events of at least Grade 3 were deemed related to study intervention. There were 3 (0.3%) serious adverse events reported; 2(0.4%) of which were from the MVC-COV1901 group while 1 (0.2%) came from the AZD1222 group; however, none of these were related to the study intervention. These SAEs included celiac disease, spontaneous abortion, and COVID-19. No death, VAED, and AE leading to study withdrawal was reported at the time the interim assessment was conducted.

### Immunogenicity outcomes

Figure 3 illustrates the rise in neutralizing antibody levels 14 days after the 2^nd^ dose of the study intervention. Among the seropositive in the PPI subset the wild-type SARS-CoV-2 neutralizing antibody GMT were 1905.6 IU/mL (95% CI 1617.98-2244.3) and 1143.4 IU/mL (95% CI 895.3-1460.2) for the MVC-COV1901 and AZD1222 groups, respectively. The GMFR from baseline were 26.0 (95% CI 19.5-34.7) and 15.0 times (95% CI 10.6-21.1) for MVC-COV1901 and AZD1222 groups, respectively. In the seropositive group, the GMT ratio between MVC-COV1901 and AZD1222 groups was 1.7 (95% CI 1.2-2.2). For the seronegative group, GMT of the MVC-COV1901 group was 434.6 IU/mL (95% CI 333.4-566.5), while that of the AZD1222 group 90.4 IU/mL (95% CI 61.1-133.9). GMFR was higher in this subsample compared to that of the seropositive group with a value of 86.2 times (95 CI% 66.4-111.9) for the MVC and 17.9 times (95% CI 12.2-26.4) for the AZD1222 groups. The GMT ratio between groups was also higher among the seronegative with the MVC-COV1901 group having a neutralizing GMT 4.8 time (95% CI 3.0-7.7) that of the AZD1222 group.

**Figure 3.**
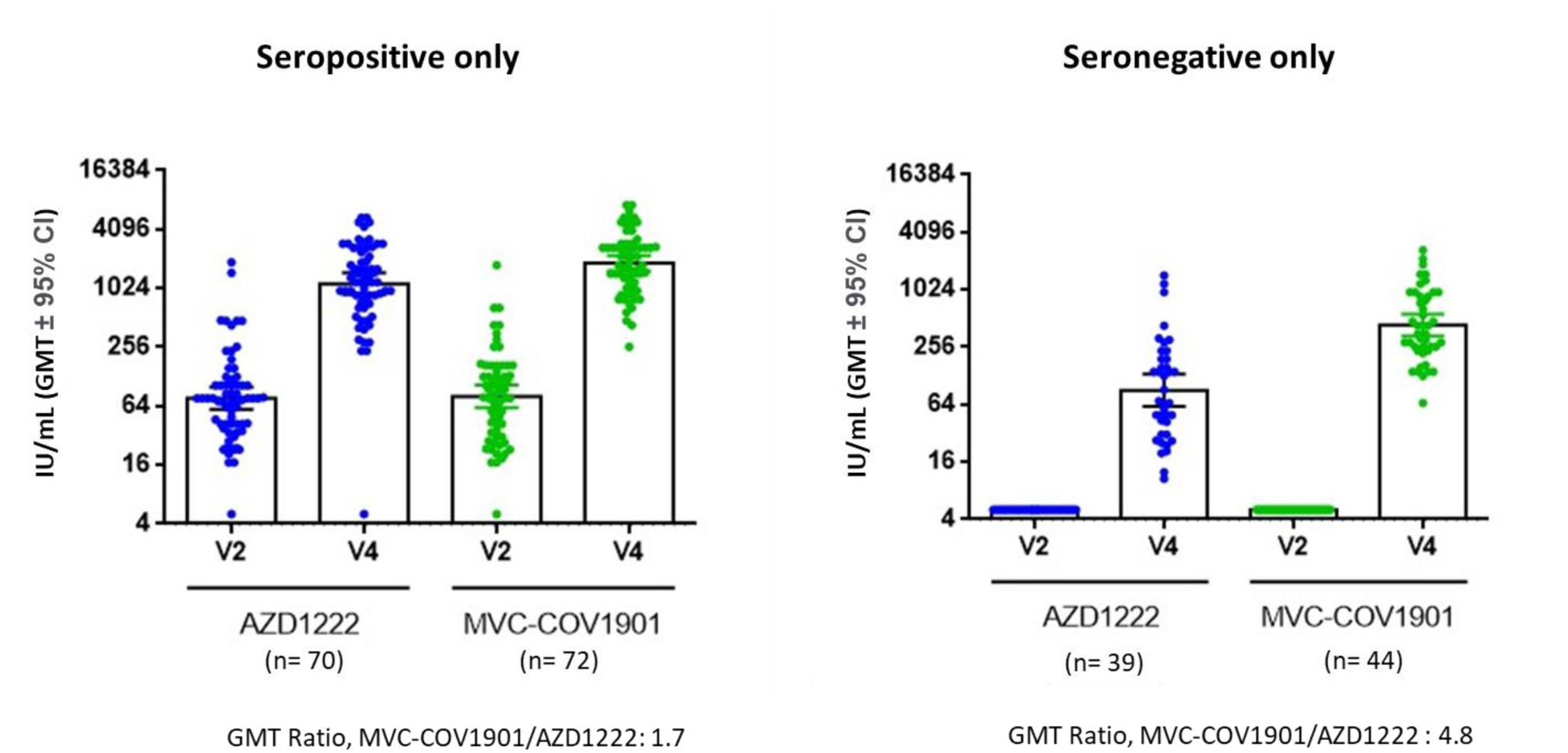
Neutralizing antibody titer in subjects immunized with two doses of either AZD1222 or MVC-COV1901 among the seropositive subsample (left) and seronegative subset. Serum samples were taken before the first vaccination (V2) and 14 days (V4) after the second dose of the study intervention and were analyzed using live SARS-CoV-2 neutralization assay. The results are shown as IU/mL with symbols indicating individual IU values and the bars indicating the GMT of each group.

Table 2 shows the seroconversion based on the wild-type SARS-CoV-2 neutralizing antibody GMTs. When considering the seronegative participants, the SCR, did not differ significantly between vaccine groups (p=0.218). The MVC-COV1901 group had a higher SCR, with only 1 participant without seroconversion. Among the seropositive, SCRs of both groups were significantly different (p=0.03). MVC-COV1901 had an SCR of 98.6 % (95% CI 95.9-100.0) while seroconversion was seen in 90.0% (95% CI 83.0-97.0) of the participants in the AZD1222 group.

**Table 2.** Seroconversion rates based on neutralizing antibody titers

| Seroconversion | AZD1222 | MVC-COV1901 | Treatment Difference<sup>1</sup>, % | p-value<sup>2</sup> |
| --- | --- | --- | --- | --- |
| Seropositive in PPI subset |  |  |  |  |
| n | 70 | 72 |  | 0.03 |
| Seroconversion, n(%) | 63 (90) | 71 (98.6) | 8.6 |  |
| 95% CI | 82.97-97.03 | 95.9-100 | 1.1-16.1 |  |
| Seronegative in PPI subset |  |  |  |  |
| n | 39 | 44 |  | 0.218 |
| Seroconversion, n(%) | 37 (94.9) | 44 (100.0) | 5.1 |  |
| 95% CI | 82.7-99.4 | 91.95-100 | -1.8-12 |  |

Abbreviation: n= no. of participants, CI= confidence interval, GMT= geometric mean titer, GMFR= geometric mean fold rise, PPI=per-protocol immunogenicity
Note: Seroconversion was defined as at least 4-fold increase of post-study intervention antibody titers from the baseline titer or from half of the lower limit of detection if undetectable at baseline.
[1] Treatment Difference was computed as, MVC-COV1901-AZD1222 and presented with the asymptotic 95% CI. In the case of small cell count (expected count less than 5), exact 95% CI was applied alternatively.
[2] P-value: Pearson's Chi-square test. In the case of small cell count (expected count less than 5), Fisher's exact test was applied alternatively

Table 3 shows the IgG titers assessed in Taiwan. As previously mentioned, testing was also done in Paraguay for local analysis and usage. Results from both Taiwan and Paraguay were positively and strongly correlated (Spearman’s r=0.80, p<0.0001). Based on data from Taiwan, the IgG GMTs in the seronegative subset of the MVC-COV1901 group increased by 239.4 times (95% CI 207.5-276.3) from baseline while that of the AZD1222 group rose by 50.8 times (95% CI 43.2-59.7). In terms of GMT ratio between MVC-COV1901 and AZD1222, results show a GMT ratio of 4.7 times (95% CI 3.8-5.9). GMFR for both groups are less when considering the seropositive subsample of the PPI subset. GMT ratio between MVC-COV1901 and AZD1222 was 1.7 times (95% CI 1.5-1.99) as measured in Taiwan. Table 4 shows that the seroconversion rate of antigen-specific immunoglobulin titers was 100% (95% CI 91.8-100) in the seronegative subsample of the MVC-COV1901 group on Day 43 after first dose. This was not significantly different from the seroconversion rate demonstrated by the AZD1222 group. Considering only the seropositive, MVC-COV1901 group had an SCR of 93.2% (95% CI 87.4-98.9) while the AZD1222 group had an SCR of 90.1% (95% CI 83.2-97.1).

**Table 3.** Geometric Mean Titers and Geometric Mean Titer Ratio of the Antigen-Specific Immunoglobulin (in BAU/mL)

| Parameter | Vaccine |  | GMT ratio;<br>(95% CI) | p-value |
| --- | --- | --- | --- | --- |
|  | AZD1222 | MVC-COV1901 |  |  |
| Seropositive |  |  |  |  |
| n | 294 | 264 |  |  |
| Baseline |  |  |  |  |
| GMT | 57.8 | 56.8 |  |  |
| 95%CI | 50.4-66.2 | 49.7-64.9 |  |  |
| Day 14 after 2nd shot |  |  |  |  |
| GMT | 1617.7 | 2812.9 | 1.7 | <0.0001 |
| 95%CI | 1460.9-1791.3 | 2575.4-3072.2 | (1.5-1.99) |  |
| GMFR | 28.0 | 49.5 |  |  |
| 95% CI | 23.6-33.2 | 42.2-58.1 |  |  |
| Seronegative |  |  |  |  |
| n | 146 | 180 |  |  |
| Baseline |  |  |  |  |
| GMT | 4.6 | 4.6 |  |  |
| 95%CI | 4.6-4.6 | 4.6-4.6 |  |  |
| Day 14 after 2nd shot |  |  |  |  |
| GMT | 231.5 | 1091.8 | 4.7 | <0.0001 |
| 95%CI | 196.7-272.4 | 945.7-1260.5 | (3.8-5.9) |  |
| GMFR | 50.8 | 239.4 |  |  |
| 95% CI | 43.2-59.7 | 207.5-276.3 |  |  |

Abbreviation: n= no. of participants, CI= confidence interval, GMT= geometric mean titer, GMFR= geometric mean fold rise
Note: [1] GMT ratio was computed as, $GMT_{MVC-COV1901}/GMT_{AZD1222}$
[2] p-value based on two-sample t test or Wilcoxon rank sum test

**Table 4.** Seroconversion rates based on Antigen-Specific Immunoglobulin Titers

| Statistics | AZD1222 | MVC-COV1901 | Treatment Difference<sup>1</sup>, % | p-value<sup>2</sup> |
| --- | --- | --- | --- | --- |
| Seropositive in PPI Subset |  |  |  |  |
| n | 71 | 73 |  | 0.5135 |
| Seroconversion, n (%) | 64 (90.1) | 68 (93.2) | 3.0 |  |
| 95% CI | (83.2-97.1) | (87.4-98.9) | (-5.9-12.0) |  |
| Seronegative in PPI Subset |  |  |  |  |
| n | 38 | 43 |  | 1 |
| Seroconversion, n (%) | 38 (100.0) | 43 (100.0) | 0 |  |
| 95% CI | (90.7-100.0) | (91.8-100.0) | (0.0-0.0) |  |

Abbreviation: n= no. of participants, CI= confidence interval, GMT= geometric mean titer, GMFR= geometric mean fold rise, PPI=per-protocol immunogenicity
Note: Seroconversion was defined as at least 4-fold increase of post-study intervention antibody titers from the baseline titer or from half of the lower limit of detection if undetectable at baseline.
[1] Treatment Difference was computed as, MVC-COV1901-AZD1222 and presented with the asymptotic 95% CI. In the case of small cell count (expected count less than 5), exact 95% CI was applied alternatively.
[2] P-value: Pearson's Chi-square test. In the case of small cell count (expected count less than 5), Fisher's exact test was applied alternatively

Assessment of IgG subclasses reveal that the pattern of IgG response in the vaccination of both AZD1222 and MVC-COV1901 is predominantly IgG1 or IgG3 (Figure 4). High levels of IgG1 and IgG3 were induced by both vaccines, with the MVC-COV1901 inducing higher IgG subclasses than the AZD1222. Among the seronegative, IgG subclass GMT ratio of the MVC-COV1901 over AZD1222 was the highest for IgG3. Minimal IgG2 and IgG4 were produced in both groups. IgG subset levels 14 days after the second dose are shown in Figure 4. The IgG1 GMTs in the seronegative is 764 BAU/mL for the MVC-COV1901 group and 208 BAU/mL for the AZD1222 group. IgG3 GMTs, on the other hand, were 608 BAU/mL and 137 BAU/mL for MVC-COV190 and AZD1222 groups, respectively. The GMT ratio between MVC-COV1901 and AZD1222 for IgG1 to IgG4 among the seronegative were 3.7, 1.4, 4.4, and 1.0, respectively.

**Figure 4.**
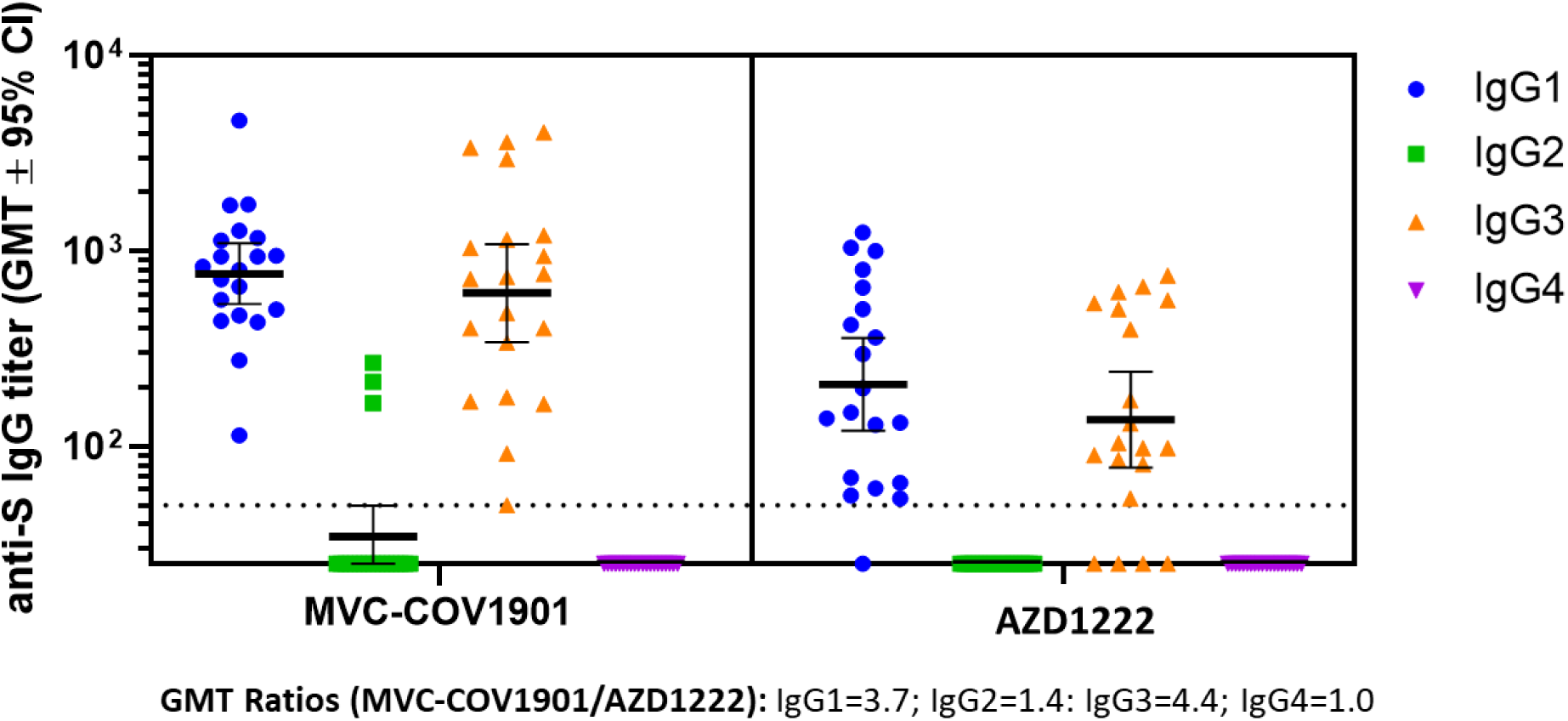
IgG subclass titers for the seronegative subsample. Depicted are geometric mean titers with 95% confidence intervals for the four subclasses of IgG.

To test the neutralizing ability against the Omicron variant, we have subjected sera collected on Day 1 and Day 43 after the first dose (i.e. Day 14 after the second dose) to pseudovirus neutralizing assay against wildtype and the Omicron variant. As shown in Figure 5, pseudovirus neutralizing antibody titers for the wildtype was higher than that of the Omicron variant among the seropositive. The baseline reciprocal inhibition dilution 50 (ID_50_) GMT for the Omicron variant was slightly higher in participants in the MVC-COV1901 group (AZD1222: 13.7 [95% CI 6.4-29.4]; MVC-COV1901: 24.3 [95% CI 8.1-73.0]) but no statistical significance was seen in both vaccine groups (p=0.3661). At Day 14 after the second dose of both vaccines, ID_50_ GMT for the neutralizing antibodies against Omicron pseudovirus was 432.0 (95% CI 76.7-2433.9) for AZD1222 and 832.2 (95% CI 389.4-1778.4) for MVC-COV1901. The MVC-COV1901 vaccine’s level of neutralizing antibodies for the Omicron pseudovirus showed a 3.0-fold (2549.7/832.2) reduction compared to the GMT of wild-type pseudovirus and was 5.2fold (2232.6/432.0) reduction for AZD1222. Between MVC-COV1901 and AZD1222, the GMT ratio of the neutralizing antibodies for the Omicron pseudovirus at day 14 after the second dose was 1.9 (95% CI 0.4-10.1). Among seronegative individuals vaccinated with MVC-COV1901, GMTs of neutralizing antibodies for the Omicron variant, were 44.5 (1323.5/29.7) times less compared to the GMTs against the wildtype pseudovirus. The GMT ratio of MVC-COV1901/AZD1222 at Day 43 for the Omicron variant was 3.0 (95% CI 1.1-8.1).

**Figure 5.**
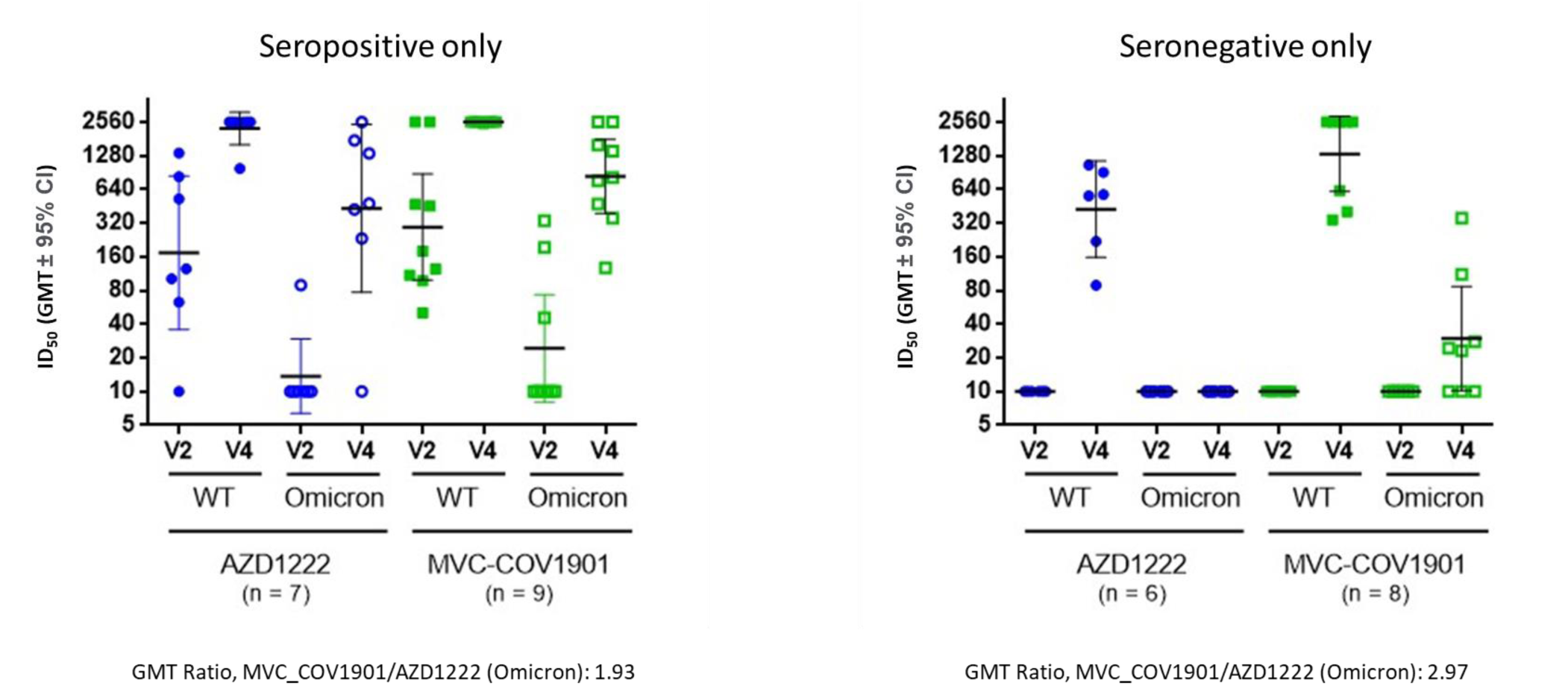
Results of pseudovirus neutralizing assay against wildtype and the Omicron variant. Serum samples taken on Day 43 after the first dose (14 days after the second dose) from 16 seropositive and 14 seronegative participants who were randomly selected. The results were presented by horizontal bars representing geometric mean titer with error bars for 95% confidence interval values.

## Discussion

In this phase III, parallel group, double-blind, randomized, active-controlled, two-arm, multi-center study, we evaluated the safety and immunogenicity of the MVC-COV1901 in adults against AZ. Initiated in October 2021, this trial met the safety and immunogenicity endpoints as stated in the protocol. Findings demonstrate the superiority of MVC-COV1901 in terms of neutralizing antibody titers and its non-inferiority in terms of resulting seroconversion rates. Neutralizing antibody GMT of MVC-COV1901 was as high as 4.8 times that of AZD1222 among the seronegative individuals. Seroconversion rate, on the other hand, was 100% among seronegative. In terms of safety. MVC-COV1901 was found to be well tolerated and have fewer safety signals compared with AZD1222 and other COVID-19 vaccines [10, 15-17].

Safety data show that adverse events are less frequent for MVC-COV1901. Reported solicited adverse events were mostly mild or moderate. However, unlike in previous studies [8,12], adverse events that are at least Grade 3 were reported for some solicited and local adverse events such as pain or tenderness, injection site pruritus, bruising, myalgia and fever. Despite this MVC-COV1901 has a good reactogenicity profile when compared with other vaccines that received an emergency use authorization [10]. A notable finding is the lower incidence of fever in the MVC-COV1901 group (10.4%) than in the AZD1222 group (19.0%). This is however, higher compared to the ones observed in the phase 2 trial conducted in Taiwan [8]. While this differential reporting of adverse events may be due to differences in age and sex composition as well as ethnicity of both populations, it is also worth noting that there are other factors which may be of significance such as differences in other social determinants, differences in experience of receiving the vaccine, and differences in the cultural context and health systems setup [18]. Unsolicited adverse events, on the other hand, were recorded on both groups. In the MVC-COV1901 group, unsolicited adverse events were reported in only 1.6% of the participants in the safety set – a rate lower compared to the reported solicited adverse events. Serious adverse events were reported by less than 1% of the participants, none of these were related to the study intervention. We suspect an underreporting of unsolicited adverse events in this clinical trial, by both participants and staff.

The phase 3 trial was conducted in Paraguay amidst the SARS-CoV-2 pandemic. Therefore, approximately 58% of the participants were seropositive at baseline. In the seronegative subset, the GMT of the wild-type neutralizing antibody in the MVC-COV1901 group increased to approximately 86 times (95% CI 66.4-111.9) from baseline. Compared to those in the AZD1222 group, the MVC-COV1901 group had a GMT that was as high as 4.8 times (95% CI 3.0-7.7) in the seronegative but in the seropositive subsample GMT ratio was 1.7 times (95% CI 1.2-2.2). This fulfills the superiority criterion set by the study (i.e. lower limit of the 95% CI must be greater than 1). In addition to this, seroconversion rates consistent with previous studies demonstrated favorable immunogenicity [8,12]. Most of the participants (99.1% in MVC-COV1901 group and 91.7% in AZD1222 group) in both vaccine groups achieved seroconversion based on wildtype neutralizing antibody on Day 14 after the second dose. Results show that the treatment difference between MVC-COV1901 and AZD1222 (i.e. MVC-COV1901 – AZD1222) was 7.4% (95% CI: 3.8 – 10.9), fulfilling the non-inferiority criterion of MVC-COV1901 to AZD1222 in SCR of neutralizing antibody. Subgroup analysis in the seronegative subsample reveal higher GMFRs (MVC: 88.8 [95% CI 68.3-115.3]; AZ: 17.9 [95%CI 12.2-26.4]). The GMT ratio between MVC-COV1901 over AZD1222 was 4.8 times (95% CI 3.0-7.7). The difference in seroconversion rates was approximately 5.1% (95% CI: -2.1 – 12.4). These also fulfill the superiority criterion for the neutralizing antibodies and non-inferiority criterion for seroconversion.

In another immunobridging study, VLA-2001, an inactivated whole virus vaccine also adjuvanted with alum and CpG 1018 adjuvant, produced a GMT ratio of 1.39 between VLA-2001 and AZD1222 [19]. Both MCV-COV1901 and VLA-2001 vaccine demonstrated superiority in neutralizing antibodies according to their set criteria. Existing literature suggest that protein subunit vaccines elicit better neutralizing antibody response compared to inactivated virus vaccines [20]. Protein subunit vaccines are purified and stably locked in the preferred pre-fusion conformation thus the vaccine is presented as a correctly folded immunogen in pure form, whereas in inactivated vaccines, the purification process may affect the spike conformations [21].

Vaccination both by MVC-COV1901 and AZD1222 induced a Th-1 skewed immune response [12,21]. AZD1222 demonstrates a Th1-biased response characterized by antibody production predominantly of IgG1 and IgG3 subclasses [22]. In similar fashion, MVC induced a robust Th-1 biased response predominated by IgG1 and IgG3. Our results illustrate that MVC induced slightly higher IgG1 and IgG3 in the seronegative population.

Levels of binding and neutralizing antibodies can be correlated and used to predict vaccine efficacy [23,24]. Reported antibody titers induced by two doses of MVC-COV1901concur with those found in previous studies [8,12] which is estimated to confer 90% vaccine efficacy against the ancestral strain [8,9]. The emergence of the other strains and particularly the Omicron strain, however, has increased ability of the virus to evade immunity, rendering two doses of currently available vaccines, largely ineffective in neutralization [25-27]. In this study, we illustrate that among seronegative participants two doses of MVC-COV1901 led to Omicron-neutralizing titers that are 44.5 times less than Wildtype-neutralizing titers. In the case of AZD1222, the reduction is more pronounced as neutralizing antibody titers against Omicron was barely detectable. As in most of the currently available vaccines, the primary regimen usually offers insufficient protection to the Omicron variant. Boosters are required to improve protection against it [27,28]. Results of this trial however, provide support to existing literature which suggests that natural immunity from previous infection offer a significant boost to protection offered by vaccination [29,30]. As aforementioned, SARS-CoV-2 was endemic in Paraguay at the time the study was conducted; hence, approximately 58% of the participants were seropositive. Because this trial started in October 2021 and because the first cases of Omicron were reported in December 2021 [31], the seropositive participants were probably infected by other SARS-CoV-2 variants. Data from seropositive individuals who got MVC-COV1901 shots show a 3.0-fold reduction in wildtype-neutralizing titers compared to Omicron neutralizing-titers viz-a-viz the 44.5-fold reduction in the seronegative. Resulting titers among the seropositive were 1.8 times higher in MVC-COV1901-vaccinated individuals compared to those vaccinated with AZD1222. A study by Nordstrom et al. [29] suggests that two-dose hybrid immunity (i.e. immunity from two doses of the vaccine plus a previous infection) was associated with 66% lower risk of reinfection than natural immunity alone with no significant attenuation up to 9 months. Two doses of hybrid immunity was also associated with a significantly lower risk of hospitalization than natural immunity. Our results, show that among those who might have been previously infected with SARS-CoV-2, the primary regimen of MVC-COV1901 induces immune response which may be sufficient against the evasive Omicron variant. In relation to existing literature, these results raise a question on issues such as the sufficiency of two doses as part of the primary vaccination regimen or the requirement of a third dose among previously infected individuals. It also highlights the idea of whether documents indicating the person’s immune status should also include a history of infection.

We consider the following limitations of the study: first, the sample size of seronegative participants was relatively small due to high local viral transmission rate in the sites at the time of the study. However, this has added to the relevance of the study in real-world settings. Second, the short duration of follow-up did not allow for the assessment of the durability of immune responses among the seropositive and seronegative participants. Third, the neutralization assay used for the Omicron variant was a pseudovirus assay which may not accurately reflect the neutralizing ability against the Omicron (BA.1) variant.

## Data Availability

Data sharing is not applicable to this article as it is an interim analysis of data from an ongoing study.

## Acknowledgements

We thank the Institute of Biomedical Sciences, Academia Sinica and Protech Pharmaservices Corporation for performing the assays. For conducting the local analysis in Paraguay, we thank Les Science Lab in Coronel Oviedo, Paraguay. We also thank the members of Medigen Vaccine Biologics Corp. in assisting manuscript editing and revision.

## Author Contribution

Concept and design: C. E. L., H.-Y. C., C. C.

Conducting the clinical trial: J.T., O.C.T., L.B., L.A.G., G.E., G.S.

Acquisition, interpretation, and analysis of data:, C. E. L., M.-Y. L., H.-Y. C., J.A.E., L.E.

Drafting of the manuscript: C. E. L., M.-Y. L., J.A.E., L.E., R.J.

