## supplementary appendix for "An Evaluation of the Safety and Immunogenicity of MVC-COV1901: Results of an interim analysis of a phase III, parallel group, randomized, double-blind, active-controlled study"

*Methods 1. Study Inclusion/Exclusion criteria*

**Inclusion Criteria**

Participants must have met all of the following inclusion criteria to be eligible for participation in this study:

1. Male or female participant aged 18 years and above at randomization.
2. Healthy adults or adults with pre-existing medical conditions who are in stable condition (obesity, endocrinological pathologies: diabetes, hyperthyroidism, hypothyroidism, etc., pneumopathies: asthma, emphysema, pulmonary fibrosis, cystic fibrosis, chronic obstructive pulmonary disease (COPD), heart diseases: arterial hypertension (HTN), arrhythmias, heart failure (HF), etc., liver diseases, renal, neurologic: angiographic cerebral vasospasm (ACV), psychiatric). A stable medical condition is defined as disease not requiring significant change in therapy or hospitalization for worsening disease 3 months before enrollment and expected to remain stable for the duration of the study.
3. Female participants:
    a. A female participant is eligible is the participant is a woman of non-childbearing potential, i.e., surgically sterilized (defined as having undergone hysterectomy and/or bilateral oophorectomy and/or bilateral salpingectomy; tubal ligation alone is not considered sufficient) or one year post-menopausal.
    b. If the participant is a woman of childbearing potential, she must agree to practice sexual abstinence or agree to use medically effective contraception from 14 days before screening to 30 days following the last administration of study intervention. Highly effective methods of contraception include:
        i. Implanted hormonal methods of contraception or placement of an intrauterine device or intrauterine hormonal-releasing system
        ii. Established use of hormonal methods (injectable, pill, patch, or ring) combined with barrier methods of contraception: condom or occlusive cap (diaphragm or cervical/vault caps) with spermicidal foam/gel/film/cream/suppository
        iii. Azoospermic partner (vasectomized or due to medical cause), provided the partner is the sole sexual partner of the female participant and the absence of sperm has been confirmed (from medical records/examination/history).
    c. Have a negative pregnancy test
4. Participant is willing and able to comply with all required study visits and follow-up required by this protocol.
5. Participant or the participant's legal representative must understand the procedures of the study and provide written informed consent.

**Exclusion Criteria:**

Participants meeting one or more of the following criteria could not be enrolled in this clinical study:

1. Pregnant or breast feeding or have plan to become pregnant within 30 days after the last administration of study intervention.
2. Employees at the investigator's site, of the Sponsor or delegate (e.g., contract research organization) who are directly involved in the conduct of the study.

**Prior/Concomitant Therapy**

3. Currently receiving or received any investigational intervention within 30 days prior to the first dose of study intervention.
4. Administered any licensed live-attenuated vaccines within 28 days or other licensed non-live-attenuated vaccines within 7 days prior to the first dose of study intervention.
5. Administered any blood product or intravenous immunoglobulin administration within 12 weeks prior to the first dose of study intervention.
6. Currently receiving or anticipate to receive concomitant immunosuppressive or immune-modifying therapy (excluding inhaled, topical skin and/or eye drop-containing corticosteroids, low-dose methotrexate, or < 2 weeks of daily receipt of prednisone less than 20 mg or equivalent) within 12 weeks prior to the first dose of study intervention.
7. Currently receiving or anticipate to receive treatment with tumor necrosis factor (TNF)-α inhibitors, e.g., infliximab, adalimumab, etanercept within 12 weeks prior to the first dose of study intervention.
8. Major surgery or any radiation therapy within 12 weeks prior to the first dose of study intervention.
9. Has received any other investigational or approved COVID-19 vaccine.

**Medical Conditions**

10. Immunosuppressive illness or immunodeficient state, including hematologic malignancy, history of solid organ, bone marrow transplantation, or asplenia.
11. A history of malignancy with potential risk for recurrence after curative treatment, or current diagnosis of or treatment for cancer (exceptions are squamous and basal cell carcinomas of the skin and treated uterine cervical carcinoma in situ, at the discretion of the investigator).
12. Bleeding disorder considered a contraindication to IM injection or phlebotomy.
13. Known SARS-CoV-2 infection in the 3 months prior to the first dose of study intervention.
14. A history of cerebral venous sinus thrombosis, heparin-induced thrombocytopenia or antiphospholipid syndrome.
15. Participant who, in the investigator's judgement, is not in a stable condition and by participating in the study could adversely affect the safety of the participant, interfere with adherence to study requirements or evaluation of any study endpoint. This may include a participant with ongoing acute diseases, severe infections, autoimmune disease, laboratory abnormality or serious medical conditions in the following systems: cardiovascular, pulmonary, hepatic, neurologic, metabolic, renal, or psychiatric.
16. A history of hypersensitivity to any vaccine or a history of allergic disease or reactions likely to be exacerbated by any component of the MVC-COV1901 or AZD1222.
17. Body (oral, rectal, or ear) temperature ≥ 38.0°C or acute illness (not including minor illnesses such as diarrhea or mild upper respiratory tract infection at the discretion of the investigator) within 2 days before the first dose of study intervention.

*Table S1-1. Summary of solicited systemic adverse events after each dosing*

| Time point, Severity | n (%) | | |
|---|---|---|---|
| | MVC-COV1901<br>N=520 | AZD-1222<br>N=510 | Total<br>N=1030 |
| **Any Solicited Systemic AEs** | | | |
| After Any Dose, All severity | 225 (49.0%) | 297 (58.2%) | 552 (53.6%) |
| After First Dose, All severity | 216 (41.5%) | 264 (51.8%) | 480 (46.6%) |
| After Second Dose, All severity | 116 (24.9%) | 106 (23.1%) | 222 (24.0%) |
| **Myalgia** | | | |
| After Any Dose, All severity | **117 (22.5%)** | **167 (32.7%)** | **284 (27.6%)** |
| Grade 1 | 110 (21.2%) | 152 (29.8%) | 262 (25.4%) |
| Grade 2 | 21 (4.0%) | 40 (7.8%) | 61 (5.9%) |
| Grade 3 | 6 (1.2%) | 4 (0.8%) | 10 (1.0%) |
| After First Dose, All severity | **86 (16.5%)** | **150 (29.4%)** | **236 (22.9%)** |
| Grade 1 | 79 (15.2%) | 132 (25.9%) | 211 (20.5%) |
| Grade 2 | 13 (2.5%) | 37 (7.3%) | 50 (4.9%) |
| Grade 3 | 1 (0.2%) | 4 (0.8%) | 5 (0.5%) |
| After Second Dose, All severity | **52 (11.2%)** | **39 (8.5%)** | **91 (9.8%)** |
| Grade 1 | 47 (10.1%) | 34 (7.4%) | 81 (8.8%) |
| Grade 2 | 10 (2.1%) | 7 (1.5%) | 17 (1.8%) |
| Grade 3 | 5 (1.1%) | 0 (0.0%) | 5 (0.5%) |
| **Malaise/Fatigue** | | | |
| After Any Dose, All severity | **107 (20.6%)** | **141 (27.6%)** | **248 (24.1%)** |
| Grade 1 | 101 (19.4%) | 126 (24.7%) | 227 (22.0%) |
| Grade 2 | 3 (0.6%) | 5 (1.0%) | 8 (0.8%) |
| Grade 3 | 3 (0.6%) | 5 (1.0%) | 8 (0.8%) |
| Grade 4 | 0 (0.0%) | 1 (0.2%) | 1 (0.1%) |
| After First Dose, All severity | **75 (14.4%)** | **115 (22.5%)** | **190 (18.4%)** |
| Grade 1 | 71 (13.7%) | 99 (19.4%) | 170 (16.5%) |
| Grade 2 | 15 (2.9%) | 28 (5.5%) | 43 (4.2%) |
| Grade 3 | 2 (0.4%) | 2 (0.4%) | 4 (0.4%) |
| Grade 4 | 0 (0.0%) | 1 (0.2%) | 1 (0.1%) |
| After Second Dose, All severity | **53 (11.4%)** | **45 (9.8%)** | **98 (10.6%)** |
| Grade 1 | 50 (10.7%) | 41 (8.9%) | 91 (9.8%) |
| Grade 2 | 12 (2.6%) | 7 (1.5%) | 19 (2.1%) |
| Grade 3 | 1 (0.2%) | 3 (0.7%) | 4 (0.4%) |
| **Headache** | | | |
| After Any Dose, All severity | **146 (28.1%)** | **192 (37.6%)** | **338 (32.8%)** |
| Grade 1 | 136 (26.2%) | 166 (32.5%) | 302 (29.3%) |

| Time point, Severity | n (%) | | |
|---|---|---|---|
| | MVC-COV1901 N=520 | AZD-1222 N=510 | Total N=1030 |
| Grade 2 | 31 (6.0%) | 45 (8.8%) | 76 (7.4%) |
| Grade 3 | 5 (1.0%) | 10 (2.0%) | 15 (1.5%) |
| After First Dose, All severity | **116 (22.3%)** | **165 (32.4%)** | **281 (27.3%)** |
| Grade 1 | 106 (20.4%) | 142 (27.8%) | 248 (24.1%) |
| Grade 2 | 21 (4.0%) | 36 (7.1%) | 57 (5.5%) |
| Grade 3 | 2 (0.4%) | 10 (2.0%) | 12 (1.2%) |
| After Second Dose, All severity | **60 (12.9%)** | **59 (12.9%)** | **119 (12.9%)** |
| Grade 1 | 53 (11.4%) | 53 (11.5%) | 106 (11.5%) |
| Grade 2 | 14 (3.0%) | 9 (2.0%) | 23 (2.5%) |
| Grade 3 | 3 (0.6%) | 1 (0.2%) | 4 (0.4%) |
| **Fever** | | | |
| After Any Dose, All severity | **54 (10.4%)** | **97 (19.0%)** | **151 (14.7%)** |
| Grade 1 | 33 (6.3%) | 73 (14.3%) | 106 (10.3%) |
| Grade 3 | 21 (4.0%) | 31 (6.1%) | 52 (5.0%) |
| After First Dose, All severity | **35 (6.7%)** | **86 (16.9%)** | **121 (11.7%)** |
| Grade 1 | 24 (4.6%) | 65 (12.7%) | 89 (8.6%) |
| Grade 3 | 11 (2.1%) | 23 (4.5%) | 34 (3.3%) |
| After Second Dose, All severity | **22 (4.7%)** | **20 (4.4%)** | **42 (4.5%)** |
| Grade 1 | 12 (2.6%) | 14 (3.1%) | 26 (2.8%) |
| Grade 3 | 10 (2.1%) | 8 (1.7%) | 18 (1.9%) |
| **Chills** | | | |
| After Any Dose, All severity | **40 (7.7%)** | **77 (15.1%)** | **117 (11.4%)** |
| Grade 1 | 38 (7.3%) | 67 (13.1%) | 105 (10.2%) |
| Grade 2 | 8 (1.5%) | 12 (2.4%) | 20 (1.9%) |
| Grade 3 | 2 (0.4%) | 3 (0.6%) | 5 (0.5%) |
| Grade 4 | 0 (0.0%) | 1 (0.2%) | 1 (0.1%) |
| After First Dose, All severity | **25 (4.8%)** | **68 (13.3%)** | **93 (9.0%)** |
| Grade 1 | 24 (4.6%) | 59 (11.6%) | 83 (8.1%) |
| Grade 2 | 6 (1.2%) | 10 (2.0%) | 16 (1.6%) |
| Grade 3 | 0 (0.0%) | 3 (0.6%) | 3 (0.3%) |
| Grade 4 | 0 (0.0%) | 1 (0.2%) | 1 (0.1%) |
| After Second Dose, All severity | **21 (4.5%)** | **15 (3.3%)** | **36 (3.9%)** |
| Grade 1 | 19 (4.1%) | 14 (3.1%) | 33 (3.6%) |
| Grade 2 | 3 (0.6%) | 2 (0.4%) | 5 (0.5%) |
| Grade 3 | 2 (0.4%) | 0 (0.0%) | 2 (0.2%) |
| **Joint Pain** | | | |
| After Any Dose, All severity | **59 (11.3%)** | **93 (18.2%)** | **152 (14.8%)** |
| Grade 1 | 53 (10.2%) | 87 (17.1%) | 140 (13.6%) |
| Grade 2 | 12 (2.3%) | 20 (3.9%) | 32 (3.1%) |

| Time point, Severity | n (%) | | |
|---|---|---|---|
| | **MVC-COV1901 N=520** | **AZD-1222 N=510** | **Total N=1030** |
| Grade 3 | 2 (0.4%) | 3 (0.6%) | 5 (0.5%) |
| Grade 4 | 0 (0.0%) | 1 (0.2%) | 1 (0.1%) |
| After First Dose, All severity | **41 (7.9%)** | **78 (15.3%)** | **119 (11.6%)** |
| Grade 1 | 40 (7.7%) | 71 (13.9%) | 111 (10.8%) |
| Grade 2 | 3 (0.6%) | 16 (3.1%) | 19 (1.8%) |
| Grade 3 | 1 (0.2%) | 3 (0.6%) | 4 (0.4%) |
| Grade 4 | 0 (0.0%) | 1 (0.2%) | 1 (0.1%) |
| After Second Dose, All severity | **23 (4.9%)** | **24 (5.2%)** | **47 (5.1%)** |
| Grade 1 | 15 (3.2%) | 22 (4.8%) | 37 (4.0%) |
| Grade 2 | 10 (2.1%) | 5 (1.1%) | 15 (1.6%) |
| Grade 3 | 1 (0.2%) | 0 (0.0%) | 1 (0.1%) |
| **Diarrhea** | | | |
| After Any Dose, All severity | **39 (7.5%)** | **44 (8.6%)** | **83 (8.1%)** |
| After First Dose, All severity | **26 (5.0%)** | **29 (5.7%)** | **55 (5.3%)** |
| After Second Dose, All severity | **14 (3.0%)** | **16 (3.5%)** | **30 (3.2%)** |
| **Nausea** | | | |
| After Any Dose, All severity | **60 (11.5%)** | **51 (10.0%)** | **111 (10.8%)** |
| After First Dose, All severity | **44 (8.5%)** | **41 (8.0%)** | **85 (8.3%)** |
| After Second Dose, All severity | **19 (4.1%)** | **16 (3.5%)** | **35 (3.8%)** |

*Table S1-2. Summary of solicited local adverse events after each dosing*

| Time point, Severity | n (%) | | |
|---|---|---|---|
| | **MVC-COV1901 N=520** | **AZD-1222 N=510** | **Total N=1030** |
| **Any Solicited Local AEs** | | | |
| After Any Dose, All severity | 209 (40.2%) | 256 (50.2%) | 465 (45.1%) |
| After First Dose, All severity | 140 (26.9%) | 202 (39.6%) | 342 (33.2%) |
| After Second Dose, All severity | 123 (26.4%) | 116 (25.3%) | 239 (25.8%) |
| **Pain/Tenderness** | | | |
| After Any Dose, All severity | **173 (33.3%)** | **215 (42.2%)** | **388 (37.7%)** |
| Grade 1 | 166 (31.9%) | 205 (40.2%) | 371 (36.0%) |
| Grade 2 | 22 (4.2%) | 42 (8.2%) | 64 (6.2%) |
| Grade 3 | 5 (1.0%) | 7 (1.4%) | 12 (1.2%) |
| Grade 4 | 0 (0.0%) | 1 (0.2%) | 1 (0.1%) |
| After First Dose, All severity | **105 (20.2%)** | **169 (33.1%)** | **274 (26.6%)** |
| Grade 1 | 96 (18.5%) | 158 (31.0%) | 254 (24.7%) |
| Grade 2 | 19 (3.7%) | 33 (6.5%) | 52 (5.0%) |
| Grade 3 | 0 (0.0%) | 6 (1.2%) | 6 (0.6%) |
| Grade 4 | 0 (0.0%) | 1 (0.2%) | 1 (0.1%) |
| After Second Dose, All severity | **100 (21.5%)** | **92 (20.0%)** | **192 (20.8%)** |
| Grade 1 | 95 (20.4%) | 86 (18.7%) | 181 (19.6%) |
| Grade 2 | 8 (1.7%) | 10 (2.2%) | 18 (1.9%) |
| Grade 3 | 5 (1.1%) | 1 (0.2%) | 6 (0.6%) |
| **Injection site pruritus** | | | |
| After Any Dose, All severity | **73 (14.0%)** | **81 (15.9%)** | **154 (15.0%)** |
| Grade 1 | 67 (12.9%) | 78 (15.3%) | 145 (14.1%) |
| Grade 2 | 9 (1.7%) | 5 (1.0%) | 14 (1.4%) |
| Grade 3 | 1 (0.2%) | 1 (0.2%) | 2 (0.2%) |
| Grade 4 | 0 (0.0%) | 1 (0.2%) | 1 (0.1%) |
| After First Dose, All severity | **60 (11.5%)** | **71 (13.9%)** | **131 (12.7%)** |
| Grade 1 | 54 (10.4%) | 68 (13.3%) | 122 (11.8%) |
| Grade 2 | 9 (1.7%) | 4 (0.8%) | 13 (1.3%) |
| Grade 3 | 1 (0.2%) | 1 (0.2%) | 2 (0.2%) |
| Grade 4 | 0 (0.0%) | 1 (0.2%) | 1 (0.1%) |
| After Second Dose, All severity | **31 (6.7%)** | **19 (4.1%)** | **50 (5.4%)** |
| Grade 1 | 31 (6.7%) | 19 (4.1%) | 50 (5.4%) |
| Grade 2 | 0 (0.0%) | 1 (0.2%) | 1 (0.1%) |
| **Erythema/Redness** | | | |
| After Any Dose, All severity | **10 (1.9%)** | **22 (4.3%)** | **32 (3.1%)** |
| Grade 1 | 8 (1.5%) | 18 (3.5%) | 26 (2.5%) |
| Grade 2 | 2 (0.4%) | 5 (1.0%) | 7 (0.7%) |
| After First Dose, All severity | **4 (0.8%)** | **14 (2.7%)** | **18 (1.7%)** |
| Grade 1 | 3 (0.6%) | 13 (2.5%) | 16 (1.6%) |
| Grade 2 | 1 (0.2%) | 3 (0.4%) | 3 (0.3%) |
| After Second Dose, All severity | **6 (1.3%)** | **8 (1.7%)** | **14 (1.5%)** |

| Time point, Severity | n (%) | | |
|---|---|---|---|
| | **MVC-COV1901 N=520** | **AZD-1222 N=510** | **Total N=1030** |
| Grade 1 | 5 (1.1%) | 5 (1.1%) | 10 (1.1%) |
| Grade 2 | 1 (0.2%) | 3 (0.7%) | 4 (0.4%) |
| **Induration/Swelling** | | | |
| After Any Dose, All severity | **10 (2.0%)** | **13 (2.5%)** | **23 (2.2%)** |
| Grade 1 | 8 (1.5%) | 11 (2.2%) | 19 (1.8%) |
| Grade 2 | 2 (0.4%) | 3 (0.6%) | 5 (0.5%) |
| After First Dose, All severity | **7 (1.4%)** | **9 (1.8%)** | **16 (1.6%)** |
| Grade 1 | 6 (1.2%) | 8 (1.6%) | 14 (1.4%) |
| Grade 2 | 1 (0.2%) | 1 (0.2%) | 2 (0.2%) |
| After Second Dose, All severity | **3 (0.6%)** | **5 (1.0%)** | **8 (0.8%)** |
| Grade 1 | 2 (0.4%) | 3 (0.6%) | 5 (0.5%) |
| Grade 2 | 1 (0.2%) | 2 (0.4%) | 3 (0.3%) |
| **Hematoma** | | | |
| After Any Dose, All severity | **42 (8.1%)** | **43 (8.4%)** | **85 (8.3%)** |
| Grade 2 | 29 (5.6%) | 32 (6.3%) | 61 (5.9%) |
| Grade 3 | 13 (2.5%) | 12 (2.4%) | 25 (2.4%) |
| After First Dose, All severity | **33 (6.3%)** | **32 (6.3%)** | **65 (6.3%)** |
| Grade 2 | 24 (4.6%) | 22 (4.3%) | 46 (4.5%) |
| Grade 3 | 9 (1.7%) | 11 (2.2%) | 20 (1.9%) |
| After Second Dose, All severity | **14 (3.0%)** | **12 (2.6%)** | **26 (2.8%)** |
| Grade 2 | 10 (2.1%) | 11 (2.4%) | 21 (2.3%) |
| Grade 3 | 4 (0.9%) | 1 (0.2%) | 5 (0.5%) |

*Table S2. Summary of unsolicited and other adverse events*

| | **MVC-COV1901 N=520** | | **AZD-1222 N=510** | | **Total N=1030** | |
|---|---|---|---|---|---|---|
| | **E** | **n (%)** | **E** | **n (%)** | **E** | **n (%)** |
| Unsolicited AEs | 11 | 8 (1.5) | 9 | 8 (1.6) | 20 | 16 (1.6) |
| Unsolicited AEs ≥ Grade 3 | 2 | 2 (0.4) | 1 | 1 (0.2) | 3 | 2 (0.3) |
| SAEs | 2 | 2 (0.4) | 1 | 1 (0.2) | 3 | 3 (0.3) |
| Related SAEs | 0 | 0 | 0 | 0 | 0 | 0 |
| AEs ≥ Grade 3 | 73 | 50 (9.6) | 95 | 69 (13.5) | 168 | 119 (11.6) |
| AESI | 1 | 1(0.2) | 0 | 0 | 1 | 1(0.1) |
| VAED | 0 | 0 | 0 | 0 | 0 | 0 |
| AEs leading to study intervention discontinuation | 2 | 2 (0.4) | 1 | 1 (0.2) | 3 | 3 (0.3) |
| AEs leading to study withdrawal | 0 | 0 | 0 | 0 | 0 | 0 |
| Death | 0 | 0 | 0 | 0 | 0 | 0 |

Abbreviation: E=no. of events, AE=adverse event, SAE= serious adverse event, AESI=adverse events of special interest, VAED= vaccine-associated enhanced disease

Note: % = percentage of participants with N as the denominator; participants might be counted in more than one category; exact 95% CIs are presented.

*Table S3. Geometric Mean Titers and Geometric Mean Titer Ratio of neutralizing antibodies (in IU/mL)*

| Parameter | Vaccine | | GMT ratio[1] (95% CI) | p-value[2] |
|---|---|---|---|---|
| | AZD1222 | MVC-COV1901 | | |
| **Seropositive** | | | | |
| n | 70 | 72 | | |
| *Baseline* | | | | |
| GMT<br>95%CI | 76.4<br>59.9-97.4 | 73.2<br>57.6-93.1 | | |
| *Day 14 after 2nd shot* | | | | |
| GMT<br>95%CI | 1143.4<br>895.3-1460.2 | 1905.6<br>1617.98-2244.3 | 1.7<br>(1.2-2.2) | <0.001 |
| GMFR<br>95% CI | 14.97<br>10.6-21.1 | 26.02<br>19.5-34.7 | | |
| **Seronegative** | | | | |
| n | 39 | 44 | | |
| *Baseline* | | | | |
| GMT<br>95%CI | 5.04<br>5.04-5.04 | 5.04<br>5.04-5.04 | | |
| *Day 14 after 2nd shot* | | | | |
| GMT<br>95%CI | 90.4<br>61.1-133.9 | 434.6<br>333.4-566.5 | 4.8<br>(3.01-7.7) | <0.0001 |
| GMFR<br>95% CI | 17.9<br>12.2-26.4 | 86.2<br>66.4-111.9 | | |

Abbreviation: n= no. of participants, CI= confidence interval, GMT= geometric mean titer, GMFR= geometric mean fold rise

Note: [1] GMT ratio was computed as, $GMT_{MVC\text{-}COV1901}/GMT_{AZD1222}$

[2] p-value based on two-sample t test or Wilcoxon rank sum test
